# Relationship of mild to moderate impairment of left ventricular ejection fraction with fatal ventricular arrhythmic events in cardiac sarcoidosis

**DOI:** 10.1101/2023.01.24.23284962

**Authors:** Yuka Akama, Yudai Fujimoto, Yuya Matsue, Daichi Maeda, Kenji Yoshioka, Taishi Dotare, Tsutomu Sunayama, Takeru Nabeta, Yoshihisa Naruse, Takeshi Kitai, Tatsunori Taniguchi, Hidekazu Tanaka, Takahiro Okumura, Yuichi Baba, Tohru Minamino

**Affiliations:** Department of Cardiovascular Biology and Medicine, Juntendo University Graduate School of Medicine, Tokyo, Japan; Department of Cardiology, Kameda Medical Center, Chiba, Japan; Department of Cardiovascular Medicine, Kitasato University School of Medicine, Sagamihara, Japan; Division of Cardiology, Internal Medicine III, Hamamatsu University School of Medicine, Hamamatsu, Japan; Department of Cardiovascular Medicine, National Cerebral and Cardiovascular Center, Osaka, Japan; Department of Cardiovascular Medicine, Osaka University Graduate School of Medicine, Osaka, Japan; Division of Cardiovascular Medicine, Department of Internal Medicine, Kobe University Graduate School of Medicine, Kobe, Japan; Department of Cardiology, Nagoya University Graduate School of Medicine, Nagoya, Japan; Department of Cardiology and Geriatrics, Kochi Medical School, Kochi University, Nankoku, Japan; Japan Agency for Medical Research and Development-Core Research for Evolutionary Medical Science and Technology (AMED-CREST), Japan Agency for Medical Research and Development, Tokyo, Japan

## Abstract

**Background:** Current guidelines recommend placing an implantable cardiac defibrillator for patients with cardiac sarcoidosis (CS) and severely impaired left ventricular ejection fraction (LVEF) of ≤35%. In this study, we aimed to explore the association between mild or moderate LVEF impairment and fatal ventricular arrhythmic event (FVAE).

**Methods:** Here, 401 patients with CS without sustained ventricular arrhythmia at diagnosis were retrospectively analyzed. The primary endpoint was an FVAE, which was defined as the combined endpoint of documented ventricular tachycardia or ventricular fibrillation and sudden cardiac death. Two cut-off points for LVEF were used; sex-specific lower threshold of normal range of LVEF, 52% for men and 54% for women, and LVEF of 35% which is used in the current guidelines.

**Results:** During a median follow-up of 3.2 years, 58 FVAEs were observed, and the 5- and 10-year estimated incidences of FVAE were 16.8% and 23.0%, respectively. All patients were classified into three groups according to LVEF: impaired LVEF group, mild to moderate impairment of LVEF group, and maintained LVEF group. Multivariable competing risk analysis showed that both the impaired LVEF group (HR: 3.02, 95% CI: 1.25–7.32) and the mild to moderate impairment of LVEF group (HR: 2.12, 95% CI: 1.02–4.40) were associated with a higher incidence of FVAE than the maintained LVEF group after adjustment for covariates.

**Conclusions:** Patients with CS are at a high risk of FVAEs, regardless of documented ventricular arrhythmia at the time of diagnosis. In patients with CS, even mild to moderate impairment of LVEF is associated with FVAEs.

**Clinical Perspective:** *What is Known:*

- Patients with cardiac sarcoidosis (CS) are at a higher risk of fatal ventricular arrhythmic event (FVAE).
- Current guidelines adopt left ventricular ejection fraction (LVEF) ≤35% as a cut-off value for Class I indication for implantable cardioverter defibrillators (ICD) implantation.

*What the Study Adds:*

- Cumulative incidence curves showed that the 5-year FVAE risk in patients with CS with preserved LVEF was 7%, which was as high as that of non-ischemic cardiomyopathy with reduced LVEF.
- For risk stratification of future fatal ventricular arrhythmic events, even milder left ventricular ejection fraction impairment, compared to that currently suggested by guidelines, needs to be considered as a risk factor in patients with cardiac sarcoidosis.
- Preventive strategies for fatal ventricular arrhythmic events and sudden cardiac death using an implantable cardiac defibrillator according to individualized risk stratification need to be developed and evaluated in clinical studies of patients with cardiac sarcoidosis.

## Introduction

Sarcoidosis is a granulomatous disease of unknown etiology that can cause multiorgan disorders in the lungs, eyes, and heart. Although cardiac sarcoidosis (CS) was previously considered to be clinically apparent in only 5% of patients with systemic sarcoidosis,(1) cardiac involvement itself has been reported in approximately 25–39% of patients in the United States (2-4) and 58% of patients in Japan.(5) Moreover, and even though pulmonary sarcoidosis is a leading cause of death, the presence of CS is strongly associated with mortality and is the second most frequent cause of death in patients with sarcoidosis.(6, 7) Given that granulomatous scars form in the myocardium and coexisting inflammation can be a substrate for macro-reentry arrhythmia in patients with CS, these patients are considered to be at a high risk for ventricular arrhythmia and sudden cardiac death (SCD).(8)

Implantable cardioverter defibrillators (ICDs) have been established as one of the most effective treatments for preventing SCD and are widely used in patients with CS.(9) The 2017 American Heart Association/American College of Cardiology/Heart Rhythm Society (AHA/ACC/HRS) Guideline for Management of Patients with Ventricular Arrhythmias and the Prevention of Sudden Cardiac Death and the 2016 Japanese Circulation Society (JCS) guidelines adopt left ventricular ejection fraction (LVEF) ≤35% as a cut-off value for Class I indication for ICD implantation.(1, 10) However, this cut-off value is derived from studies examining the efficacy of ICD in patients with heart failure with reduced ejection fraction rather than CS, and the optimal cut-off value for LVEF is still to be determined in terms of risk stratification of SCD. Indeed, previous studies have shown that patients with CS whose LVEF was >35% were also associated with a higher risk of ventricular tachycardia and ventricular fibrillation (VT/VF), and SCD.(11, 12) The aim of this study was to examine the association between mild to moderate impairment of LVEF and fatal ventricular arrhythmias and SCD in patients with CS.

## Methods

### Study subjects

The ILLUMINATE-CS (ILLUstration of the Management and prognosIs of JapaNese PATiEnts with Cardiac Sarcoidosis) was designed as a multicenter retrospective registry to evaluate the clinical characteristics and outcomes of patients with CS.(13) The inclusion criterion for this study was a diagnosis of CS according to either the JCS criteria proposed in 2016 or the HRS 2014 consensus statement.(1, 14) In contrast, the exclusion criterion was the patient’s refusal to be enrolled after notification that they were registered in this registry. Among the 33 participating hospitals, 21 were university hospitals and 12 were non-university teaching hospitals. This study complied with the Declaration of Helsinki and Japanese Ethical Guidelines for Medical and Health Research Involving Human Subjects. The study protocol was approved by the ethics committee of each participating hospital, and the requirement for informed consent was waived because of the retrospective nature of the study. Study information, including the objectives, inclusion and exclusion criteria, and names of participating hospitals, was published in the publicly available University Hospital Information Network (UMIN000034974).

### Data collection and outcomes

Baseline characteristics, including age, sex, clinical comorbidities, blood test data, and findings of cardiovascular imaging tests, were obtained during the initial diagnostic process for CS. The presenting manifestations were determined as all documented manifestations at the time of CS diagnosis. The baseline was defined as the time at which a patient was diagnosed with CS by fulfilling either the JCS or HRS criteria. Past medical history, arrhythmias, and conduction disorders at baseline were the main presenting manifestations at the time of diagnosis. For instance, if a patient’s first cardiac manifestation was VT/VF, leading to the diagnosis of CS after evaluation, then this was recorded as “history of VT/VF: yes.”

All patients were stratified by LVEF using two cut-off values: sex-specific lower threshold of normal range of LVEF proposed by the American Society of Echocardiography and the European Association of Cardiovascular Imaging (i.e. 52% for men and 54% for women)(15); and LVEF of 35%, which is used for class I recommendation for ICD implantation in the current guidelines.(10) Based on the following cut-off values, patients were classified into three groups: impaired LVEF group (≤35%), mild to moderate impairment of LVEF group (>35% and <52% for men, and <35% and <54% for women), and maintained LVEF group (≥52% for men and ≥54% for women).

Cardiac accumulation on late gadolinium enhancement (LGE) on cardiac magnetic resonance (CMR) imaging was determined from reports provided by experts at each institution. We used the AHA 17-segment model to evaluate the distribution and semi-quantify the segments of the myocardium showing hyperenhancement on CMR-LGE.(16)

As the primary endpoint, this study assessed fatal ventricular arrhythmia events (FVAEs), which were defined as SCDs coupled with documented VF, sustained VT lasting for >30 seconds, or appropriate ICD therapy. SCD and heart failure hospitalization were defined according to the definitions recently proposed by the Heart Failure Collaboratory and Academic Research Consortium.(17)

### Statistical analysis

Normally distributed continuous variables are expressed as mean ± standard deviation, whereas non-normally distributed variables are presented as the median and interquartile range (IQR). Categorical variables are expressed as numbers and percentages. A one-way analysis of variance or the Kruskal–Wallis test was used to evaluate group differences for continuous variables, and the chi-square test or the Fisher’s exact test for dichotomous variables, as appropriate. When necessary, the variables were transformed for further analyses. Multiple imputations were performed to consider missing data in the derivation cohort, and survival analyses were performed after 20 imputed datasets without missing data were created using a chained-equations procedure. Cumulative incidence curves for an FVAE from the time of CS diagnosis (time zero) were generated by Fine–Gray competing risk regression analysis, with death that was not a consequence of an FVAE as a competing risk. Univariate Fine–Gray competing risk regression analysis was used to identify variables potentially associated with the primary outcome. The optimal cut-off for LVEF predictive of FVAEs was determined using the Youden index in receiver operating characteristic curve analysis. Variables with a *P*-value of <0.1 on univariate analysis were considered for inclusion in a multivariable analysis.

## Results

Among the 512 patients registered in the ILLUMINATE-CS registry, we excluded 111 patients because of missing data on LVEF (n = 13), history of VT/VF (n = 24), or previously documented VT/VF (n = 74) at the time of diagnosis. A total of 401 patients were included in the final cohort. Among them, 257 (64.1 %) had histological evidence of sarcoidosis in cardiac or extracardiac tissues, and 39 (9.7%) were diagnosed with CS directly by endomyocardial biopsy. According to the diagnostic criteria, 369 patients met the JCS criteria, and 251 met the HRS criteria. During the median follow-up period of 3.2 (IQR: 1.7–5.8) years, 58 FVAEs were observed. The 5- and 10-year estimated incidence rates of FVAE were 16.8% and 23.0%, respectively. In all patients, the mean LVEF was 50.7% (IQR: 37.5–63%) at the time of CS diagnosis.

The baseline characteristics of the patients stratified by LVEF groups are shown in Table 1. Patients with more impaired LVEF were associated with a history of admission for heart failure, non-sustained ventricular tachycardia, more severe heart failure symptoms, high brain natriuretic peptide values, impaired renal function, and angiotensin-converting enzyme inhibitor/angiotensin receptor blocker and beta-blocker use. In addition, more patients with impaired LVEF were treated with a defibrillator. Only 248 out of the 401 patients included in our analysis underwent CMR, and the percentage of patients presenting with LGE at any segment and the number of segments showing LGE enhancement were higher in those with more impaired LVEF. Cumulative incidence curves for FVAE stratified by LVEF categories are shown in Figure 1. The 5- and 10-year estimated incidence rates of FVAE were 7.0% and 15.1% for the maintained LVEF group, 20.0% and 28.5% for the mild to moderate impairment of LVEF group, and 30.2% and 33.4% for the impaired LVEF group. Both the impaired LVEF group and the mild to moderate impairment of LVEF group were associated with a higher incidence of FVAE than that of the maintained LVEF group (Fine–Gray: *P* < 0.001). This association did not change even after analyzing only the patients who met the HRS criteria and JCS criteria (Figure S1).

**Table 1.** Patient characteristics stratified by LVEF categories.

| Variables | Impaired LVEF<br>group<br>N = 80 | Mild to moderate<br>impairment of LVEF<br>group<br>N = 137 | Maintained<br>LVEF group<br>N = 184 | P-value |
| --- | --- | --- | --- | --- |
| Age (years) | 61.7 ± 10.7 | 61.8 ± 11.0 | 61.2 ± 12.2 | 0.878 |
| Male sex (%) | 28 (35.0) | 54 (39.4) | 52 (28.3) | 0.105 |
| Past medical history (%) |  |  |  |  |
| Hypertension | 28 (35.9) | 61 (44.5) | 67 (36.6) | 0.286 |
| Diabetes | 21 (26.6) | 45 (33.1) | 51 (27.9) | 0.497 |
| HF admission | 42 (52.5) | 26 (19.0) | 10 (5.4) | <0.001 |
| NSVT | 18 (22.5) | 28 (20.7) | 20 (11.2) | 0.024 |
| High-degree AV block | 33 (41.2) | 58 (42.3) | 85 (46.4) | 0.656 |
| Atrial fibrillation | 8 (10.0) | 19 (14.2) | 13 (7.1) | 0.118 |
| NYHA III/IV (%) | 25 (31.6) | 13 (9.8) | 12 (6.9) | <0.001 |
| Laboratory data at baseline |  |  |  |  |
| BNP (pg/mL) | 364.7 [164.1–<br>592.8] | 138.9 [63.3–375.5] | 64.7 [28.7–<br>137.1] | <0.001 |
| Cre (mg/dL) | 0.81 [0.69–1.04] | 0.78 [0.66–0.99] | 0.74 [0.63–<br>0.88] | 0.005 |
| eGFR (mL/min/1.73 m<sup>2</sup>) | 79.8±22.5 | 84.9±23.9 | 88.0±18.0 | 0.019 |
| LVEF (%) | 28.0 [22.8–32.0] | 44.0 [40.0–48.9] | 64.0 [58.1–<br>69.7] | <0.001 |
| Findings on CMR in patients who underwent CMR* |  |  |  |  |
| Presence of LGE | 43 (95.6) | 78 (94.0) | 100 (86.2) | 0.081 |
| Number of segments with LGE hyperenhancement | 5 [3–8] | 4 [2–5] | 3 [1–4] | <0.001 |
| Medication at baseline |  |  |  |  |
| ACE-i | 35 (44.9) | 44 (32.4) | 18 (9.8) | <0.001 |
| ARB | 21 (26.9) | 41 (30.1) | 38 (20.8) | 0.15 |
| Beta blocker | 55 (70.5) | 66 (48.5) | 27 (14.8) | <0.001 |
| Amiodarone | 10 (12.8) | 4 (2.9) | 6 (3.3) | 0.002 |
| Steroid (at any timepoint during f/u) | 67 (83.8) | 127 (92.7) | 163 (88.6) | 0.122 |
| Devices at baseline |  |  |  |  |
| Pacemaker/CRT-P | 20 (25.6) | 39 (29.1) | 53 (29.1) | 0.831 |
| ICD/CRT-D | 12 (16.0) | 6 (4.5) | 6 (3.4) | <0.001 |

\*Only 248 patients underwent CMR imaging.
ACE-I, angiotensin-converting enzyme inhibitor; ARB, angiotensin receptor blocker; AV, atrioventricular; BNP, brain natriuretic peptide; CMR, cardiac magnetic resonance; Cre, creatinine; CRT-D, cardiac resynchronization therapy defibrillator; CRT-P, cardiac resynchronization therapy pacemaker; eGFR, estimated glomerular filtration rate; f/u: follow-up; HF, heart failure; ICD, implantable cardioverter defibrillator; LGE, late gadolinium enhancement; LVEF, left ventricular ejection fraction; NSVT, non-sustained ventricular tachycardia.

**Figure 1.**
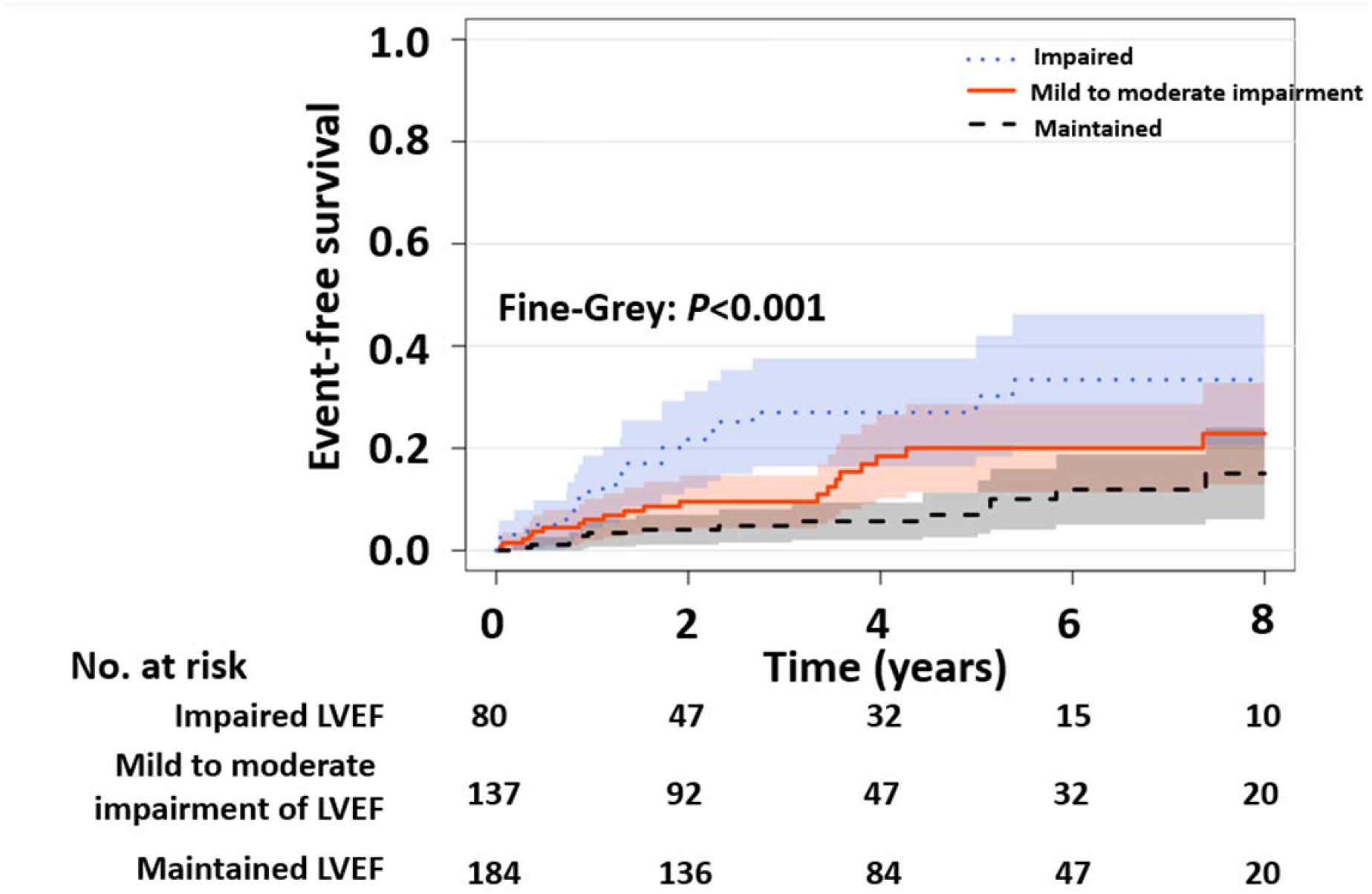
Cumulative incidence curves for fatal ventricular arrhythmic events (FVAEs) stratified by impaired, mild to moderate impairment, and maintained left ventricular ejection fraction (LVEF).

The univariate and multivariate competing risk analyses results are shown in Table 2. Although several factors were identified as potentially associated with FVAE in the univariate analysis, including the number of segments showing LGE enhancement, only impaired LVEF (HR: 3.02, 95% CI: 1.25–7.32) and mild to moderate impairment of LVEF (HR: 2.12, 95% CI: 1.02–4.40) were found to be associated with FVAE in the multivariable Fine–Gray model. Moreover, there was no significant difference in the association between the impaired LVEF group and mild to moderate impairment of LVEF group in terms of association with FVAE (*P* = 0.290). ROC curves were constructed to test the discriminating performances of LVEF and to determine the optimal cut-off for LVEF predictive FVAEs stratified by sex. The best cut-off points were LVEF <48% for men and LVEF <54% for women, respectively.

**Table 2.**
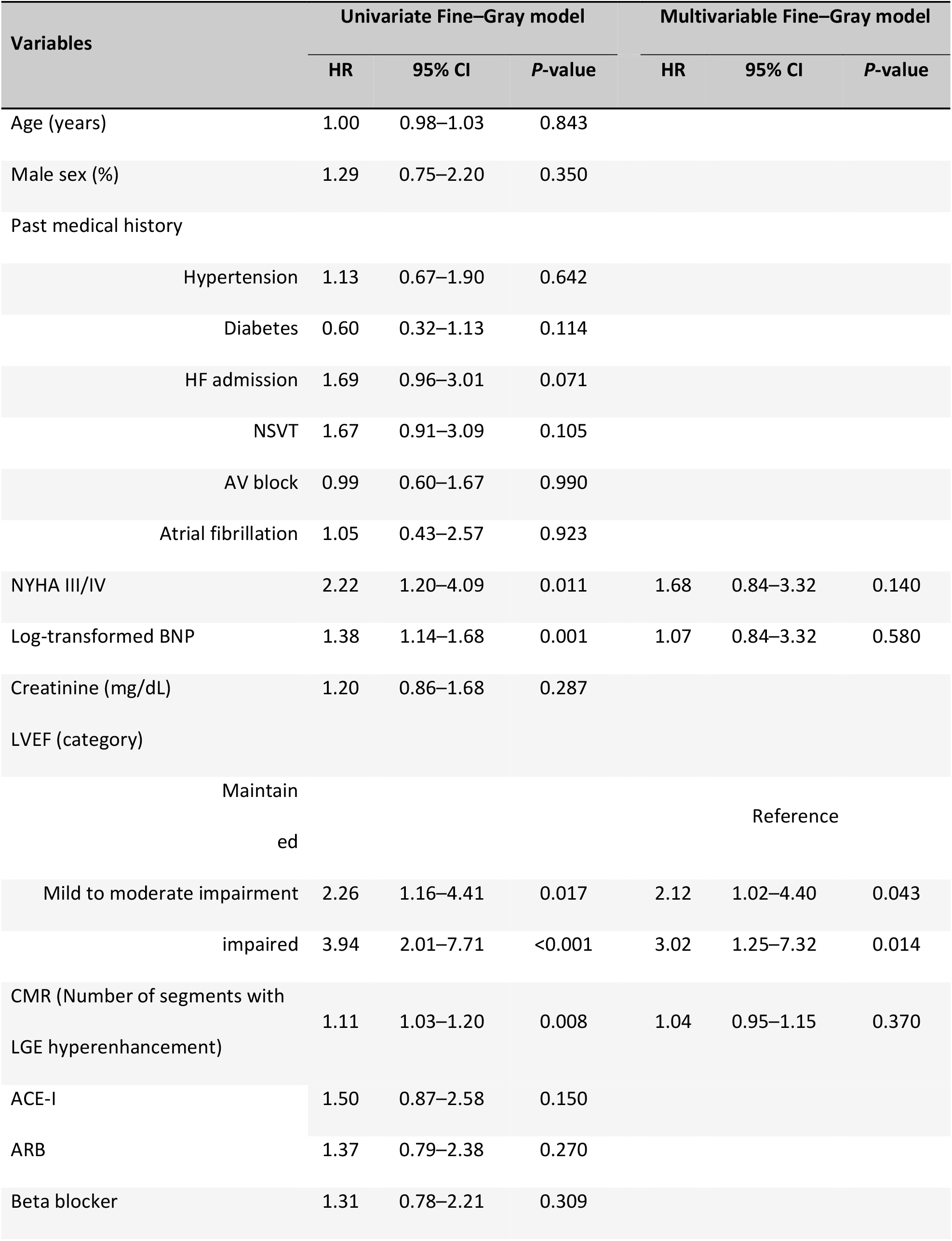
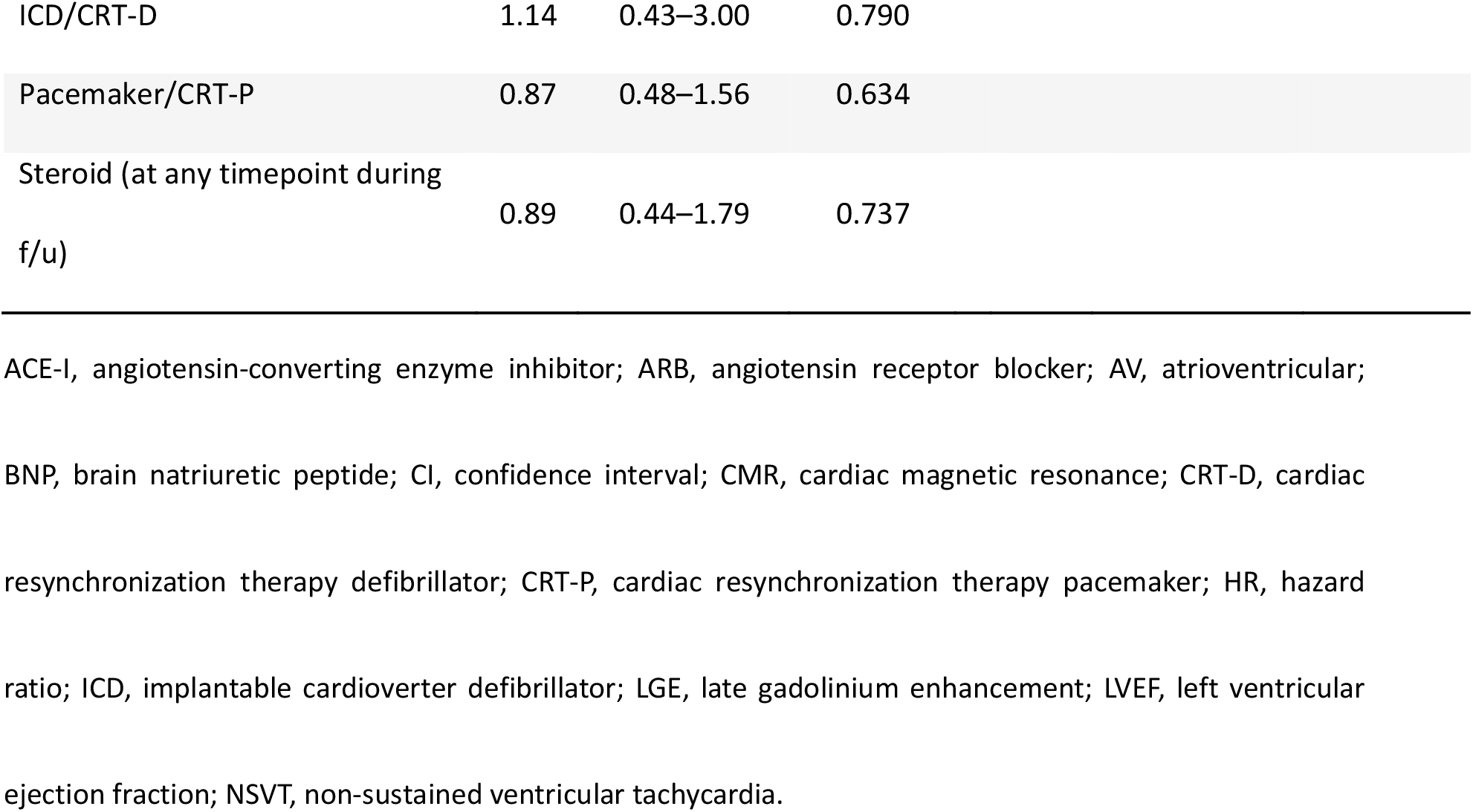
Univariate and multivariable Fine–Gray competing risk models.

| Variables | Univariate Fine–Gray model |  |  | Multivariable Fine–Gray model |  |  |
| --- | --- | --- | --- | --- | --- | --- |
|  | HR | 95% CI | P-value | HR | 95% CI | P-value |
| Age (years) | 1.00 | 0.98–1.03 | 0.843 |  |  |  |
| Male sex (%) | 1.29 | 0.75–2.20 | 0.350 |  |  |  |
| Past medical history |  |  |  |  |  |  |
| Hypertension | 1.13 | 0.67–1.90 | 0.642 |  |  |  |
| Diabetes | 0.60 | 0.32–1.13 | 0.114 |  |  |  |
| HF admission | 1.69 | 0.96–3.01 | 0.071 |  |  |  |
| NSVT | 1.67 | 0.91–3.09 | 0.105 |  |  |  |
| AV block | 0.99 | 0.60–1.67 | 0.990 |  |  |  |
| Atrial fibrillation | 1.05 | 0.43–2.57 | 0.923 |  |  |  |
| NYHA III/IV | 2.22 | 1.20–4.09 | 0.011 | 1.68 | 0.84–3.32 | 0.140 |
| Log-transformed BNP | 1.38 | 1.14–1.68 | 0.001 | 1.07 | 0.84–3.32 | 0.580 |
| Creatinine (mg/dL) | 1.20 | 0.86–1.68 | 0.287 |  |  |  |
| LVEF (category) |  |  |  |  |  |  |
| Maintained |  |  |  | Reference |  |  |
| Mild to moderate impairment | 2.26 | 1.16–4.41 | 0.017 | 2.12 | 1.02–4.40 | 0.043 |
| Impaired | 3.94 | 2.01–7.71 | <0.001 | 3.02 | 1.25–7.32 | 0.014 |
| CMR (Number of segments with LGE hyperenhancement) | 1.11 | 1.03–1.20 | 0.008 | 1.04 | 0.95–1.15 | 0.370 |
| ACE-I | 1.50 | 0.87–2.58 | 0.150 |  |  |  |
| ARB | 1.37 | 0.79–2.38 | 0.270 |  |  |  |
| Beta blocker | 1.31 | 0.78–2.21 | 0.309 |  |  |  |

|  |  |  |  |
| --- | --- | --- | --- |
| ICD/CRT-D | 1.14 | 0.43–3.00 | 0.790 |
| Pacemaker/CRT-P | 0.87 | 0.48–1.56 | 0.634 |
| Steroid (at any timepoint during<br>f/u) | 0.89 | 0.44–1.79 | 0.737 |

ACE-I, angiotensin-converting enzyme inhibitor; ARB, angiotensin receptor blocker; AV, atrioventricular; BNP, brain natriuretic peptide; CI, confidence interval; CMR, cardiac magnetic resonance; CRT-D, cardiac resynchronization therapy defibrillator; CRT-P, cardiac resynchronization therapy pacemaker; HR, hazard ratio; ICD, implantable cardioverter defibrillator; LGE, late gadolinium enhancement; LVEF, left ventricular ejection fraction; NSVT, non-sustained ventricular tachycardia.

## Discussion

In this study, we analyzed patients diagnosed with CS according to the latest guidelines and found that the 5- and 10-year estimated incidence rates of FVAE were 16.8% and 23.0%, respectively. The incidence of FVAE for at 5 and 10 years in those with maintained LVEF were as high as 7.0% and 15.1%, respectively. We further found that not only LVEF below the cut-off value proposed by the current guidelines (as a Class I indication for ICD implantation for the prevention of SCD) but also mild to moderate impairment of LVEF were independently associated with further increased risk of FVAE in patients without documented ventricular arrhythmia at the time of admission. This association was independent of the extent of the scar, which was measured as the hyperenhancement of CMR-LGE. Our study results suggest the possibility that even a mild decline in LVEF needs to be recognized as a risk factor for future FVAE in patients with CS.

Considering the fact that patients with CS are at a higher risk of FVAE,(11, 18) risk stratification and appropriate recommendations for ICD are essential. Despite some differences between the guidelines, LVEF remains one of the most important indications in all guidelines and consensus documents. Numerous observational studies, including the present study, have clearly shown that low LVEF is strongly associated with FVAE in patients with CS; therefore, low LVEF is the only criterion used for the class I recommendation for ICD implantation in patients with CS without documented ventricular arrhythmia. However, although all current guidelines and consensus documents use a 35% LVEF cut-off value, this was not derived from studies investigating patients with CS and has instead been mainly derived from randomized controlled trials that showed the prognostic implications of ICD in patients with heart failure with reduced ejection fraction. This implies that even though the association between LVEF equal to or below 35% and high FVAE risk has also been well validated in patients with CS, it does not necessarily suggest that this LVEF cut-off value is optimal for risk stratification of FVAE in this population. Indeed, a few studies with limited sample size have shown that even a mild reduction in LVEF is associated with ventricular arrhythmia and/or SCD. A study of 112 patients with CS treated with ICD found that LVEF of <55% was a risk factor for the need for appropriate ICD therapy for ventricular arrhythmias.(19) This is in line with our analysis results suggesting that the best cut-off points to predict FVAE were LVEF <48% for men and LVEF <54% for women, which are close to the sex-specific lower threshold of the normal range of LVEF proposed by the American Society of Echocardiography and the European Association of Cardiovascular Imaging. In another study investigating 235 patients with CS and implanted ICD, more than one-third of the patients (85 of 235) who received appropriate ICD therapy had an LVEF of >35%.(18) Moreover, Betensky et al. reported that seven out of 17 patients who received appropriate ICD therapy had an LVEF of >35%.(11) As a consequence, recently updated ESC guidelines on cardiac pacing and cardiac resynchronization therapy have adopted a higher LVEF cut-off value, and recommend that patients with CS who require cardiac pacing and have a LVEF of <50% should be considered for a cardiac resynchronization therapy defibrillator as opposed to a pacemaker (class IIa recommendation and level of evidence C).(20) Based on the large number of events and the sizable number of patients diagnosed with CS in accordance with the most recent guidelines, the present study supports the validity of this recommendation. Additionally, we focused only on the indication for ICD as primary prevention, unlike other studies that included patients with documented ventricular arrhythmia who were indicated for ICD regardless of LVEF.(11, 18, 19)

The importance of CMR-LGE as a risk factor for FVAE in patients with sarcoidosis has been previously reported. A meta-analysis showed that CMR-LGE is a powerful predictor of all-cause death or arrhythmogenic events (ventricular arrhythmia, ICD shock, and SCD) in patients with known or suspected CS.(21) In another meta-analysis involving 471 patients with suspected or diagnosed (cardiac or extracardiac) sarcoidosis, the presence of LGE was associated with an 8.6-fold higher risk of ventricular arrhythmias.(22) A similar study included 290 patients with biopsy-proven sarcoidosis regardless of cardiac involvement or known or suspected CS; in that study, CMR-LGE in >5.7% of the whole myocardium was suggested to be associated with a composite endpoint of ventricular arrhythmia or SCD.(23) Additionally, these findings are consistent with our study results, which demonstrate an association between CMR-LGE hyperenhancement and FVAE risk in the univariate model. According to the results of our analysis LVEF, rather than the number of segments with CMR-LGE hyperenhancement, was associated with FVAE. However, this result should be carefully interpreted as it does not necessarily provide a definitive conclusion regarding the superiority of the modality to evaluate the risk of FVAE in patients with CS, given the limited number of patients who underwent CMR compared to echocardiography. Additionally, we did not perform more detailed measurements of CMR data. Nevertheless, the significance of our study results may not be undermined in any significant way, given the fact that echocardiography has several advantages over CMR regarding the risk stratification of FVAE in patients with CS. First, echocardiography is superior to CMR in terms of cost-effectiveness and accessibility. Second, not all patients with cardiac implantable electrical devices, which is a very common treatment option for patients with CS, can undergo CMR. Third, CMR-LGE was contraindicated in patients with severe renal impairment. Lastly, repetitive evaluation of cardiac function using echocardiography is easier and less costly than follow-up with CMR. This is particularly important, considering that serial imaging evaluations after treatment with immunosuppressive agents are recommended when evaluating the indications for ICD.(1, 14)

Our results showed that low LVEF measured by echocardiography was associated with FVAE regardless of the extent of the CMR-LGE hyperenhancement area. We believe that echocardiography can play a vital role in predicting the risk of future FVAE, despite the fact that CMR may also provide valuable information.

It should be also noted that our study results reinforced the fact that CS is a disease associated with high risk of FVAE even if LVEF is maintained. A Finnish cohort study including 398 patients with myocardial or extracardiac biopsy-proven CS demonstrated that the 5-year risk of SCD was approximately 5% even without the current guidelines’ ICD indications.(24) Our competing risk regression analysis showed that the estimated 5-year risk of FVAE was as high as 7% even in the maintained LVEF group. These two study results are very consistently showing that the FVAE risk in patients with CS with preserved LVEF is as high as non-ischemic cardiomyopathy with reduced LVEF, which is demonstrated to be 6.3%.(25) There has been no consensus on the cut-off value of the 5-year risk of SCD to determine ICD implantation for patients with CS, but the European guideline for hypertrophic cardiomyopathy indicates that ICD should be considered if the estimated 5-year risk of SCD is ≥6% and may be considered if the risk is 4% to 6%.(26) Taking this into account, our study results demonstrated the importance of impaired LVEF as a risk factors for FVAE, while at the same time indicating that LVEF alone does not adequately assess the risk. Further research is warranted to better risk-stratify for sudden cardiac death in patients with CS.

### Limitations

This study had some limitations. First, as this was a retrospective study, there was a potential for sampling bias and missing data. Second, although we tried to standardize data quality as much as possible, including the findings on advanced cardiovascular imaging and our inclusion of experienced and board-certified cardiologists to collect data, there could be disparities in the accuracy of data owing to the nature of the multicenter retrospective study design. For example, the extent of LGE on CMR might not be assessed accurately because the protocol of CMR might differ among the institutions. Third, we only used LVEF measured at the time of diagnosis and did not consider changes in LVEF, which can be observed after treatment with immunosuppressive agents; however, as the impact of immunosuppressive agents on LVEF remains unclear and is unpredictable,(27) the optimal timing of evaluating LVEF to risk stratification for ICD implantation is unknown. Therefore, this limitation does not seriously affect the significance of our study results. Fourth, this study included patients diagnosed with CS at participating institutions over a period of 17 years; therefore, a limited number of patients had advanced cardiovascular imaging data available, including CMR, as some were enrolled before these imaging techniques were developed. Moreover, even though we performed a semi-quantitative analysis for the finding of CMR, a more advanced analysis using appropriate software was not conducted. Fifth, it is probable that not all FVAEs were accurately recorded, and the FVAE rate was, therefore, underestimated, particularly in patients without cardiac implantable electrical devices. However, it is important to note that patients with impaired LVEF and mild to moderate impairment of LVEF experienced similar FVAE events, although the number of patients with cardiac implantable electrical devices in the impaired LVEF group was three times higher than that in the mild to moderate impairment group. Finally, we used two different diagnostic criteria to determine eligibility, and patients without histological evidence were included in this study according to the Japanese Circulation Society guidelines. However, we demonstrated that our findings did not change significantly when only patients satisfying the HRS criteria were analyzed.

In conclusion, we found that patients with CS are at a high risk of FVAE, even if no ventricular arrhythmia is documented at the time of diagnosis. Even milder impairment of LVEF above the cut-off value suggested in the current guidelines is associated with FVAEs in patients with CS. Therefore, further studies are required to develop optimal risk stratification strategies for patients with CS.

## Data Availability

Data not available due to ethical restrictions.

## Acknowledgements

None

## Conflict of interest statement

Dr. Yuya Matsue received an honorarium from Otsuka Pharmaceutical Co and Novartis Japan. Dr. Takahiro Okumura received honoraria from Ono Yakuhin, Otsuka, Novartis, and Astrazeneca, as well as research grants from Ono Yakuhin, Amgen Astellas, Pfizer, Alnylam, and Alexion (not in connection with the submitted work). Other authors have nothing to declare.

### Abbreviations

CS: cardiac sarcoidosis
SCD: sudden cardiac death
ICD: implantable cardioverter defibrillators
AHA/ACC/HRS: American Heart Association/American College of Cardiology/Heart Rhythm Society
JCS: Japanese Circulation Society
LVEF: left ventricular ejection fraction
VT/VF: ventricular tachycardia and ventricular fibrillation
LGE: late gadolinium enhancement
CMR: cardiac magnetic resonance
IQR: interquartile range

## Supplemental Material

Figure S1. Cumulative incidence curves for fatal ventricular arrhythmic events (FVAEs) stratified by impaired, mild to moderate impairment, and maintained left ventricular ejection fraction (LVEF) in patients who met (A) the Heart Rhythm Society (HRS) diagnostic criteria and (B) Japanese Circulation Society criteria.

